# Validation of a novel fluorescent lateral flow assay for rapid qualitative and quantitative assessment of total anti-SARS-CoV-2 S-RBD binding antibody units (BAU) from plasma or fingerstick whole-blood of COVID-19 vaccinees

**DOI:** 10.1101/2022.01.04.22268754

**Authors:** Nadin Younes, Duaa W. Al-Sadeq, Farah M. Shurrab, Hadeel T. Zidan, Haissam Abou-Saleh, Bushra Y. Abo Halawa, Fatima M. AlHamaydeh, Amira E. Elsharafi, Hanin I. Daas, Swapna Thomas, Sahar Aboalmaaly, Afra Al Farsi, Reeham Al-Buainain, Samar Ataelmannan, Jiji Paul, Amana Salih Al Saadi, Hadi M. Yassine, Amin F. Majdalawieh, Ahmed Ismail, Laith J. Abu-Raddad, Gheyath K. Nasrallah

## Abstract

**Background:** Limited commercial LFA assays are available to provide a reliable quantitative measurement of the total binding antibody units (BAU/mL) against the receptor-binding domain of the SARS-CoV-2 spike protein (S-RBD).

**Aim:** To evaluate the performance of FinecareTM2019-nCoV S-RBD LFA and its fluorescent reader (FinecareTM-FIA Meter) against the following reference methods (i) The FDA-approved Genscript surrogate virus-neutralizing assay (sVNT), and (ii) three highly performing automated immunoassays: BioMérieux VIDAS®3, Ortho VITROS®, and Mindray CL-900i®.

**Methods:** Plasma from 488 vaccinees were tested by all aforementioned assays. Fingerstick whole-blood samples from 156 vaccinees were also tested by FinecareTM.

**Results and conclusions:** FinecareTM showed 100% specificity as none of the pre-pandemic samples tested positive. Equivalent FinecareTM results were observed among the samples taken from fingerstick or plasma (Pearson correlation *r*=0.9, p<0.0001), suggesting that fingerstick samples are sufficient to quantitate the S-RBD BAU/mL. A moderate correlation was observed between FinecareTM and sVNT (*r=*0.5, p<0.0001), indicating that FinecareTM can be used for rapid prediction of the neutralization antibody post-vaccination. FinecareTM BAU results showed strong correlation with VIDAS®3 (*r=*0.6, p<0.0001), and moderate correlation with VITROS® (*r*=0.5, p<0.0001), and CL-900i® (*r*=0.4, p<0.0001), suggesting that FinecareTM be used as a surrogate for the advanced automated assays to measure S-RBD BAU/mL.

## Introduction

Serological immunoassays are rapidly emerging with varying degrees of sensitivity and specificity (1, 2). These include enzyme-linked immunosorbent assays (ELISAs), chemiluminescence assays (CLIA), electrofluorescent (ELFA), and lateral flow assays (LFAs). Most serological assays measure the binding antibodies (BA) response against specific SARS-CoV-2 antigens. Measuring binding antibodies is insufficient for assessing protective immunity since these antibodies (Abs) do not play significant roles in virus neutralization (3, 4). neutralizing antibodies (nAbs) are more indicative of protective immunity due to their ability not only to bind S-RBD, but also to block viral entry to the host cells (3, 5–8). Therefore, neutralization assays remain the gold standard for measuring the nAbs titer against SARS-CoV-2 (9). Nevertheless, nAb assays require high-level biosafety laboratories, highly trained personnel and often take several days to complete (9). To avoid such limitations, Genscript Biotech Corporation developed a reliable surrogate virus-neutralizing assay (known as sVNT or cPass) to measure the nAb. The cPass Genscript assay is now recommended by the World Health Organization (WHO) and also received the full USA FDA approval to be used as a reference assay for quantitative measurement of the nAb (10).

One of the major challenges for the commercially available serology assays is that their binding antibody results are provided in arbitrary units per milliliters (ARU/mL). Thus, the results between assays are highly variable, although they target the same SARS-CoV-2 antigen, usually the S-protein. To minimize the discrepancies between serological assays, the WHO introduced an international standard to harmonize the antibodies immune response assessment and recommended reporting the assay results for binding activity in binding antibody unit per milliliter (BAU/mL) instead of arbitrary ARU/mL. (11). One of the best ways to identify a reliable assay that can be used as a surrogate assay to assess the quantity of neutralizing Abs post-vaccination is by performing correlation studies between these assays and a reference neutralization assay.

LFAs are attractive for small or non-laboratory settings and population surveillance. They are affordable and rely on easily accessible specimens such as fingerstick whole blood, and provide the result within minutes. According to the literature, none of the few commercially available fluorescent LFAs that provide quantitative measurement of SARS-CoV-2 (in BAU/mL) to the S-RBD protein was validated. The FinecareTM2019-nCoV S-RBD Antibody Test is a fluorescence immunoassay used along FinecareTM FIA Meters (reader) (Model No.: FS-113) for quantitative detection of S-RBD BAU in human fingerstick whole blood, venipuncture whole blood, and serum or plasma specimens (12). For more information about the assay principle, see **Figure S1**. In this study, we aimed to evaluate the performance of FinecareTM2019-nCoV by comparing its results with (i) the Genescript a surrogate virus-neutralizing test (sVNT), and (ii) three highly performing commercially available automated anti-SARS-CoV-2 immunoassays; Mindray CL-900i® SARS-CoV-2 S-RBD IgG and BioMérieux VIDAS®3, which measure BAU for S-RBD (13, 14), and Ortho VITROS®, which measures total BAU to the S1 spike protein subunit (13, 14).

## 2. Methods

### 2.1 Samples collection and ethical approval

Participants who received two BNT162b2 or mRNA-1273 vaccine doses were eligible for inclusion. A total of 488 EDTA whole-blood samples (5.0 mL) were collected between April and October 2021 from staff and students at Qatar University, the largest national university in Qatar. Plasma was separated from whole venous blood and stored at −80°C until performing the test. Demographic information and information on the previous infection with SARS-CoV-2 were collected through a self-administered questionnaire. Out of the total samples, 156 participants in the vaccinated group performed the FinecareTM test with fingerstick whole blood. The study was reviewed and approved by the Institutional Review Board at Qatar University (QU-IRB 1537-FBA/21). All analyses were conducted according to the ethical standards of the Declaration of Helsinkiof the World Medical Association (WMA).

### 2.2 FincareFinecareTM anti-SARS-CoV-2 S-RBD total antibodies test

#### 2.2.1 Fingerstick samples

Briefly, the area of the fingertip was lanced with an alcohol pad. Then, the skin was punctured using a sterile lancet, and 20 μL of fingerstick blood was collected with the capillary sampler. The specimen was squeezed out into the detection buffer tube. After that, 75 μL of sample mixture was loaded into the sample well and was inserted into the Test Cartridge holder of FinecareTM FIA Meters. The reaction time is 15 minutes. The results unit is displayed as a relative fluorescence unit (RFU, AU/mL). To obtain the results in BAU/mL, the AU/mL values were multiplied by the WHO standard (20) that is provided by the manufacturer.

#### 2.2.2 Plasma samples

According to the standard phlebotomy procedure, venipuncture whole blood specimens were collected from each participant in EDTA tubes. Then, plasma was separated from blood immediately, and the test was performed as described above in 2.2.1.

### 2.3 Commercially available serological assays

Samples were analyzed using three commercially available automated anti-SARS-CoV-2 immunoassays assays: Mindray CL-900i® SARS-CoV-2 IgG (Cat. No. SARS-CoV-2 S-RBD IgG122, Mindray, Shenzhen, China), VIDAS®3 SARS-CoV-2 IgG (Cat. No. 423834, BioMérieux, Marcy-l’Étoile, France), and Ortho VITROS® anti-SARS-CoV-2 total Ab (Ortho Clinical Diagnostics, USA). Each assay was performed according to the manufacturer’s instructions. The characteristics of the assays, including detection method, targeted antigens, sample volume, result interpretation, reported sensitivity and specificity, and WHO conversion factor to BAU/mL are summarized in **Table 1**.

**Table 1.** Characteristics of the automated analyzers used in this study.

| Manfac<br>turer | Autom<br>ated<br>Analyz<br>er | Detc<br>tion<br>Meth<br>od | Targ<br>eted<br>Antig<br>en | Samp<br>le<br>Volum<br>e | Result<br>Interpre<br>tation | Repor<br>ted<br>Sensit<br>ivity | Repor<br>ted<br>Specif<br>icity | WHO<br>Conve<br>rsion<br>Factor | Refer<br>ence |
| --- | --- | --- | --- | --- | --- | --- | --- | --- | --- |
| Mindray<br>Bio-Medical<br>Electron<br>ics Co.,<br>Ltd. | Mindra<br>y CL-900i®<br>SARS-CoV-2<br>S-RBD<br>IgG | CLIA<br>* | S-RBD | 10 µL<br>(this<br>volum<br>e does<br>not<br>includ<br>e the<br>dead<br>volum<br>e) | <10<br>AU/mL:<br>Negative<br>≥10<br>AU/mL:<br>Positive | 100%<br>(≥15<br>days) | 94.9% | 1.15 | (15,<br>16) |
| BioMéri<br>eux<br>Diagnos<br>tics | VIDA<br>S®3<br>SARS-CoV-2<br>IgG | ELFA<br>* | S-RBD | 100<br>µL<br>(inclu<br>ding<br>the<br>dead<br>volum<br>e) | <1<br>AU/mL:<br>Negative<br>≥1<br>AU/mL:<br>Positive | 100%<br>(≥15<br>days) | 98.5% | 20.3 | (17) |
| Ortho<br>Clinical<br>Diagnos<br>tics | Ortho<br>VITR<br>OS®<br>anti-SARS-CoV-2<br>Total<br>Ab | CLIA | S (S1<br>subun<br>it) | 20 µL<br>(this<br>volum<br>e does<br>not<br>includ<br>e the<br>dead<br>volum<br>e) | <1.0 S/C:<br>Negative<br>≥1.0 S/C:<br>Positive | NR | 100% | 17.8 | (18) |

NR: Not reported
\*CLIA, Chemiluminescence Immunoassay; ELFA, Enzyme-Linked Fluourescent Assay; S: Spike Protein; N: Nucleocapsid Protein; S1: Subunit of the Spike Protein. NA: Not Applicable.

### 2.4 cPass Genscript sVNT

A SARS-CoV-2 surrogate virus neutralization test (sVNT) (Cat. No. L00847, GenScript Biotech, NJ, USA) was utilized as a reference test for detecting nAbs against SARS-CoV-2. The test was conducted following the manufacturer’s specifications. Briefly, samples and controls (provided within the kit) were diluted 1:10 with the sample dilution buffer provided with the kit. Then, 125□ μL of sample/control was mixed in a 1:1 ratio with HRP-RBD solution and incubated at 37°C for 30 minutes. After that, 100□µl of each sample/control was added to the hACE2 coated plate in duplicates. The plate was sealed and incubated for 15 minutes at 37°C. Wells were then washed 4x with 200□µl of the wash solution provided. 100□µl of the TMB solution was added per well and the plate was incubated in the dark for 15 minutes at room temperature. Finally, 50 μL of stop solution was added to each well to quench the reaction, and absorbance was read at 450□nm. % inhibition of ≥30% was considered positive, and percent inhibition of <30% was considered negative. Conversion tool for cPass % inhibition to IU/mL of the WHO International Standard (NIBSC code 20/136) was calculated as established in this (10).

### 2.5 Statistical method

Correlation and linear regression analysis between FinecareTM and each commercial serological immunoassay and the sVNT were performed. Pearson correlation coefficient (r) was calculated. For absolute values of Pearson’s r, 0–0.19 is denoted as very weak, 0.2–0.39 as weak, 0.40–0.59 as moderate, 0.6–0.79 as strong, and 0.8–1 as very strong correlation (19). Concordance analysis between FinecareTM and the three automated assays was conducted, which includes the overall percent agreement (OPA), positive percent agreement (PPA), and negative percent agreement (NPA), accuracy/efficiency as well as Cohen’s Kappa statistic. Cohen’s Kappa measure is a robust metric that estimates the level of agreement between two diagnostic tests. A Kappa value <0.40 denotes poor agreement, 0.40–0.59 denotes fair agreement, 0.60–0.74 denotes good agreement, and ≥0.75 denotes excellent agreement (20). The significance level was indicated at 5%, and a 95% confidence interval (CI) was reported for each metric. All statistical analysis was performed using GraphPad Prism software (Version 8.2.1. San Diego, CA, USA).

## 3. Results

### 3.1. FinecareTM is a very specific and sensitive assay

To assess the specificity of the of the FinecareTM, we tested 100 pre-pandemic plasma samples using FinecareTM as well as sVNT as a reference method. None of the tested samples were positive by any of the two assay, indicating 100% specificity of the FinecareTM. Regarding the sensitivity, 156 plasma samples were tested by the sVNT and the FinecareTM. 155 samples were positive by sVNT and FinecareTM. However, only one sample tested negative with sVNT but positive with FinecareTM indicating 99.8% sensitivity of the FincareTM. Intrestingly this sample tested negative when fingerstick whole blood sample were tested. The discripency between the reslts of plasma and fingerstick could be due to a technical error between the two different technichains who performed the test.

### 3.2. FinecareTM fingerstick and plasma results are comparable

We assessed the performance of FinecareTM using fingerstick whole blood samples. A total of 156 participants provided informed consent to receive a fingerstick and have venous blood drawn to generate matched plasma samples. These samples were tested using the FinecareTM and results were compared as shown in **Figure 1**. A very strong correlation was observed (*r*=0.9, P<0.0001) between fingerstick and venous plasma samples. Similarly, linear regression analysis showed that constructed model could strongly predict the dependent variable (R^2^=0.8, p<0.0001). Most importantly, the interpretation concordance was 100%, demonstrating equivalency between fingerstick and venous plasma.

**Figure 1.**
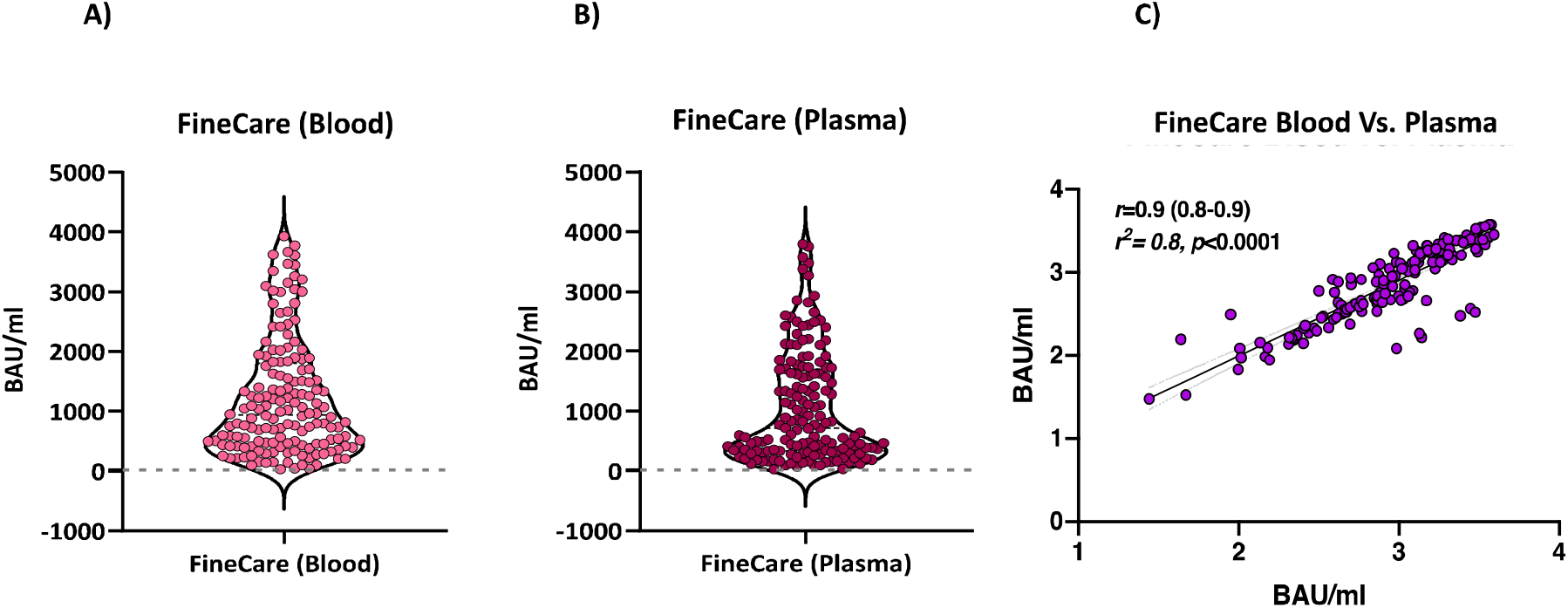
**A)** Point distribution of fingerstick whole blood samples. **B)** Point distribution of plasma samples. **C)** Correlation and linear regression analysis between matched fingerstick whole blood sample and plasma sample using FinecareTM (n□=□156). Pearson correlation coefficient (*r*), coefficient of determination (R^2^), and *p*-value are shown.

### 3.3 Moderate correlation between FinecareTM and sVNT

We evaluated the performance of FinecareTM in comparison to sVNT. Moderate significant correlation between FinecareTM and sVNT with *r-*value of 0.5 (p□<□0.001) was shown in **Figure 2**. Most importantly, the interpretation concordance was 99.4% as only one sample tested negative with sVNT but positive with FinecareTM.

**Figure 2.**
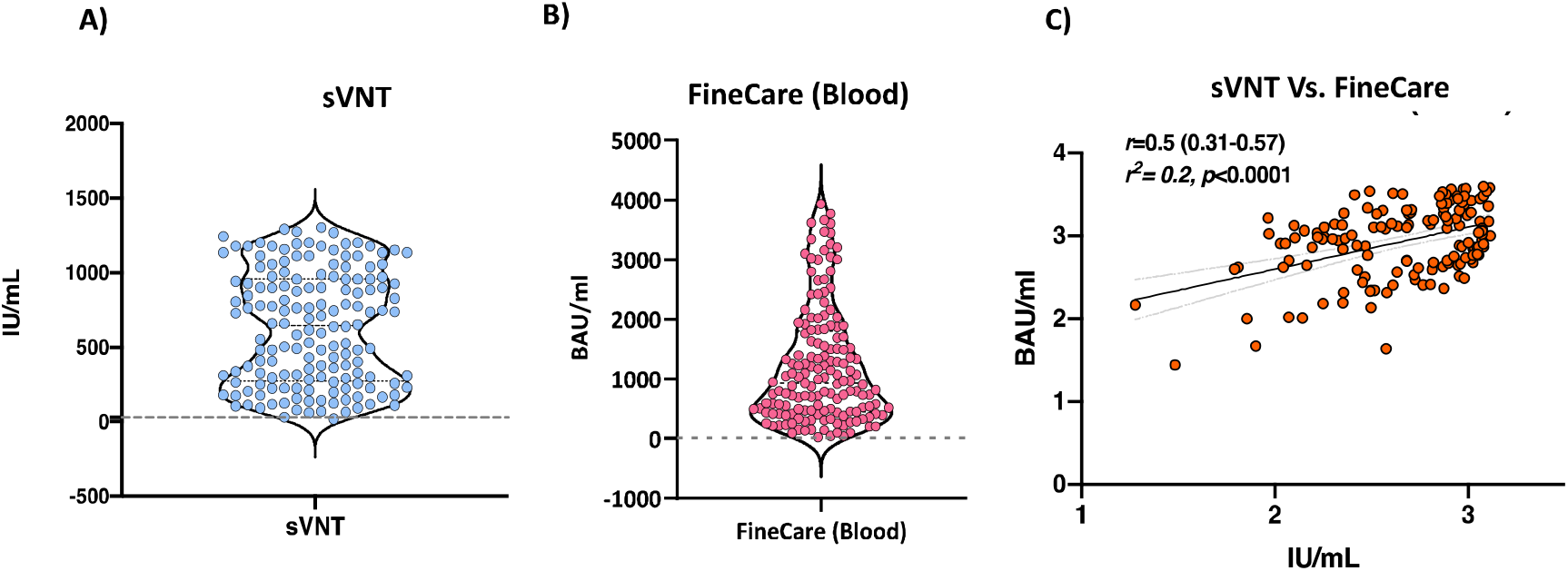
**A)** Point distribution of sVNT. **B)** Point distribution of FinecareTM. **C)** Correlation and linear regression analysis between FinecareTM and the surrogate virus neutralization test (sVNT). Pearson correlation coefficient (*r*), coefficient of determination (R^2^), and *p*-value are shown.

### 3.4 Moderate to strong correlation between FinecareTM and three commercially available automated anti-SARS-CoV-2 immunoassays

The correlation and linear regression analysis between the readings obtained from FinecareTM and each automated immunoassay are illustrated in **Figure 3**. Pearson’s correlation coefficients (r) showed a statistically significant positive correlation for FinecareTM with all three automated assays (p<0.001). The strongest correlation coefficient was shown with VIDAS®3 (*r*=0.6, **Figure 3A**), followed by VITROS® (*r*=0.5, **Figure 3C**), and CL-900i® (*r*=0.4, **Figure 3B**). Linear regression analysis showed that all constructed models could statistically significantly predict the dependent variable [Fine care (BAU/mL)]. The best regression model fitting the data was shown by VIDAS®3 (R^2^ = 0.3), followed by VITROS® (R^2^ = 0.3) and CL-900i® (R^2^ = 0.2).

**Figure 3.**
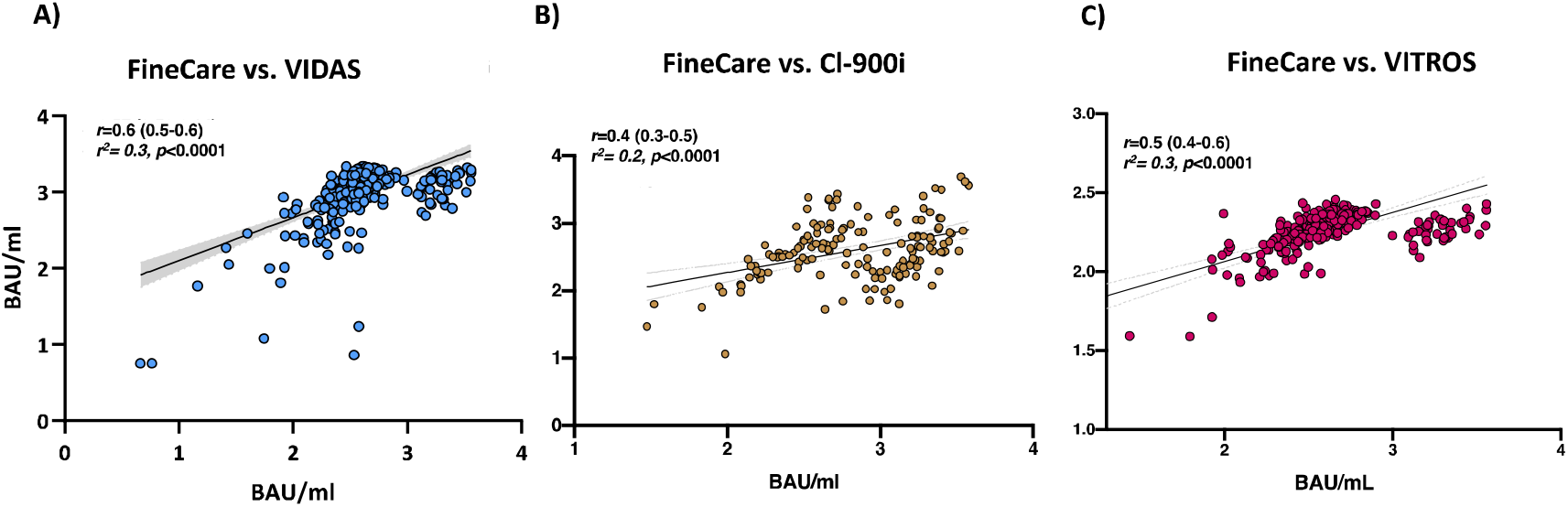
Correlation and linear regression analysis between FinecareTM and **A)** VIDAS®3 **B)** CL-900i® **C)** VITROS®. Pearson correlation coefficient (*r*), coefficient of determination (R^2^), and *p*-value are shown.

### 3.5. Excellent agreement between FinecareTM and the three automated anti-SARS-CoV-2 immunoassays

The tests’ agreements were studied in a pairwise fashion applying inter-rater agreement statistics; (Cohen’s Kappa statistic, k) (**Table 2**). The OPA, PPA, NPA were 100% between FinecareTM and the three automated immunoassays. In addition, Cohen’s Kappa statistic denoted excellent agreement between FinecareTM and the three automated anti-SARS-CoV-2 immunoassays (κ= 1.00). Most importantly, the interpretation concordance was 100%, demonstrating equivalency between FinecareTM and the three automated anti-SARS-CoV-2 immunoassays.

**Table 2.** Concordance assessment between FinecareTM and the three automated anti-SARS-CoV-2 immunoassays (CL-900i®, VIDAS®3, and VITROS®).

| Test | Compared to | Overall Percent Agreement (OPA) | Positive Percent Agreement (PPA) | Negative Percent Agreement (NPA) | Accuracy/Efficiency | Cohen's Kappa Statistic |
| --- | --- | --- | --- | --- | --- | --- |
|  |  | % (95% CI) | % (95% CI) | % (95% CI) | % (95% CI) | k (95% CI) |
| Finecare<sup>TM</sup> | VIDAS<sup>®</sup>3 | 100 (98.90-100) | 100 (98.90-100) | 100 (2.5-100.00) | 100 (98.90-100.00) | 1.00 (0.40-0.86) |
|  | CL-900i<sup>®</sup> | 100 (99.25-100.00) | 100 (99.25-100.00) | 100 (15.81-100) | 100 (99.25-100.00) | 1.00 (0.52 to 0.96) |
|  | VITROS<sup>®</sup> | 100 (98.85-100) | 100 (98.84-100) | 100 (2.5-100.00) | 100 (98.85-100.00) | 1.00 (0.46 - 0.95) |

## 4. Discussion

Despite this rapid progress in vaccines development, it is unlikely that COVID-19 will be eradicated in the near future. At best, COVID-19 infections can be brought under control to the point where life can return to “pre-COVID” normality (21). A key part of this approach will be the deployment of high throughput testing to determine who is immune and the duration of immune response (21). The rapid LFA-POC antibody tests are simple, cheap, and fast. They do not require qualified personnel for interpretation and could be done in non-laboratory settings. The usefulness of rapid tests for detection of SARS-CoV-2 Abs has been questioned regarding their inferiority compared to other serology assays. This is because none of the few commercially available LFAs provide quantitative measurement of SARS-CoV-2 (in BAU/mL) to the S-RBD. The FinecareTM2019-nCoV S-RBD Antibody Test used along FinecareTM reader is the first quantitative LFA for the detection of S-RBD BAU in human fingerstick whole blood, venipuncture whole blood, and serum or plasma specimens (12). WHO International Standard (BAU/mL) is essential for facilitating the standardization of SARS-CoV-2 serological methods and allow for comparison and harmonization of datasets across laboratories. This is critical for determining the antibody levels that are needed for efficacious vaccines and protection from emerging variants such as Omicron (22). Recent studies have shown that two-dose COVID-19 vaccine regimen do not induce enough neutralizing antibodies against the Omicron variant (22).

One of the major advantages of FinecareTM is using fingerstick whole blood samples and obtaining quantitative results within 15 minutes. Although Abs are more stable in serum/plasma samples, the whole blood samples are more convenient to use. Therefore, we assessed the performance of FinecareTM using fingerstick whole blood samples compared to venous plasma samples. A very strong correlation was observed (*r*=0.9, P<0.0001) between fingerstick and venous plasma samples. In addition, the interpretation concordance was 100%, demonstrating equivalency between fingerstick whole blood and venous plasma samples. Our results confirmed the feasibility of using fingerstick whole blood for SARS-CoV-2 IgG detection and showed excellent concordance with plasma values using FinecareTM assay. Collection of fingerstick whole blood samples in Microtainer tubes is quick and easy. It does not require a phlebotomist, making it an attractive alternative to venepuncture for use in SARS-CoV-2 drive-through testing sites. The simplicity of performing the LFA with whole blood also eliminates the need for centrifugation and plasma separation steps, reducing the cost and complexity of obtaining a result.

FinecareTM, which targets S-RBD antibodies, showed a moderate degree of correlation (*r*=0.5, p□<0.001) with the sVNT and demonstrated 100% specificity (data not shown). Most importantly, the interpretation concordance between FinecareTM and sVNT reached 99.4%. S-RBD plays an essential role in viral entry into the cells suggesting neutralizing properties and immunity against SARS-CoV-2 (5–8). Studies have shown that serology assays that detect Abs against the S1 subunit or the RBD alone strongly correlate with neutralization activity (23–27). According to McAndrews et al., approximately 86% of individuals positive for S-RBD–binding Abs exhibited neutralizing capacity (3). Whereas Abs directed against the Nucleocapsid protein (NP) are only binding Abs with no neutralizing activity (3, 28).

The correlation and linear regression analysis between the readings obtained from FinecareTM and each automated anti-SARS-CoV-2 immunoassay was evaluated. Each assay was selected for previously determined performance (13, 14), ease-of-use characteristics (automated serology) and availability. Pearson’s correlation coefficients (r) showed a statistically significant positive correlation between FinecareTM and all three automated assays (p < 0.001). The strongest correlation was shown with VIDAS®3 (*r*=0.6), followed by VITROS® (*r*=0.5), and CL-900i® (*r*=0.4). The best correlation with VIDAS®3 was expected as both assays are based on fluorescence immunoassay technology and solely target Abs against S-RBD. Cohen’s Kappa statistic denoted excellent agreement between FinecareTM and the three automated anti-SARS-CoV-2 immunoassays. Interestingly, the interpretation concordance was 100%, demonstrating equivalency between FinecareTM and the three automated immunoassays. The excellent concordance between the POC FinecareTM and the automated serological assay makes it an attractive alternative for serological assays that do not require laboratory settings. The ability to rapidly, accurately, and affordably determine seroprevalence in a population will be an important tool in the growing arsenal of SARS-CoV-2 diagnostic testing and assessment of antibody response post-vaccination, particularly in resource-limited areas.

In conclusion, SARS-CoV-2 continues to spread and threatens to disrupt human activity for years to come. Fortunately, we already have vaccines that effectively reduce disease and are now being rolled out in many countries but countering that many countries have low vaccine coverage and ongoing high rates of infection, raising the potential for immune-mediated selection escape variants. FinecareTM should contribute to more widespread access to immunity testing for many populations, including those in resource-poor areas with limited access to laboratories and equipment for more conventional virus neutralization tests or ELISA-based inhibition assays.

## Data Availability

All data produced in the present work are contained in the manuscript

## 5. Author contributions

Conceptualization, GKN and LJA.; methodology, FMS, DWA, FMH, AES, BYH, ST, SAB, AA, RA, SA, JP, and AS; software, FMS, DWA, FMH, AES, BYH, SAB, AA, RA, SA, JP, and AS.; validation, FMS, DWA, FMH, AES, BYH, SAB, AA, RA, SA, JP, and AS.; formal analysis, GKN, FMS, HTZ, and NY investigation, GKN and LJA.; resources, GKN, LJA, and AA.; data curation, GKN, FMS, HTZ, and NY.; writing—original draft preparation, NY, DWA, and GKN; writing—review and editing, HID, HA, AFM and GKN.; visualization GKN, FMS, HTZ, and NY.; supervision, HA, AA, HMY, and GKN.; project administration, GKN and LJA.; funding, GKN, LJA, and AA. All authors have read and agreed to the published version of the manuscript.

## 8. Ethical approval

The study was reviewed and approved by the Institutional Review Board at Qatar University (QU-IRB 1537-FBA/21). All analyses were conducted according to the ethical standards of the Declaration of Helsinkiof the World Medical Association (WMA).

---

**Figure S1.**
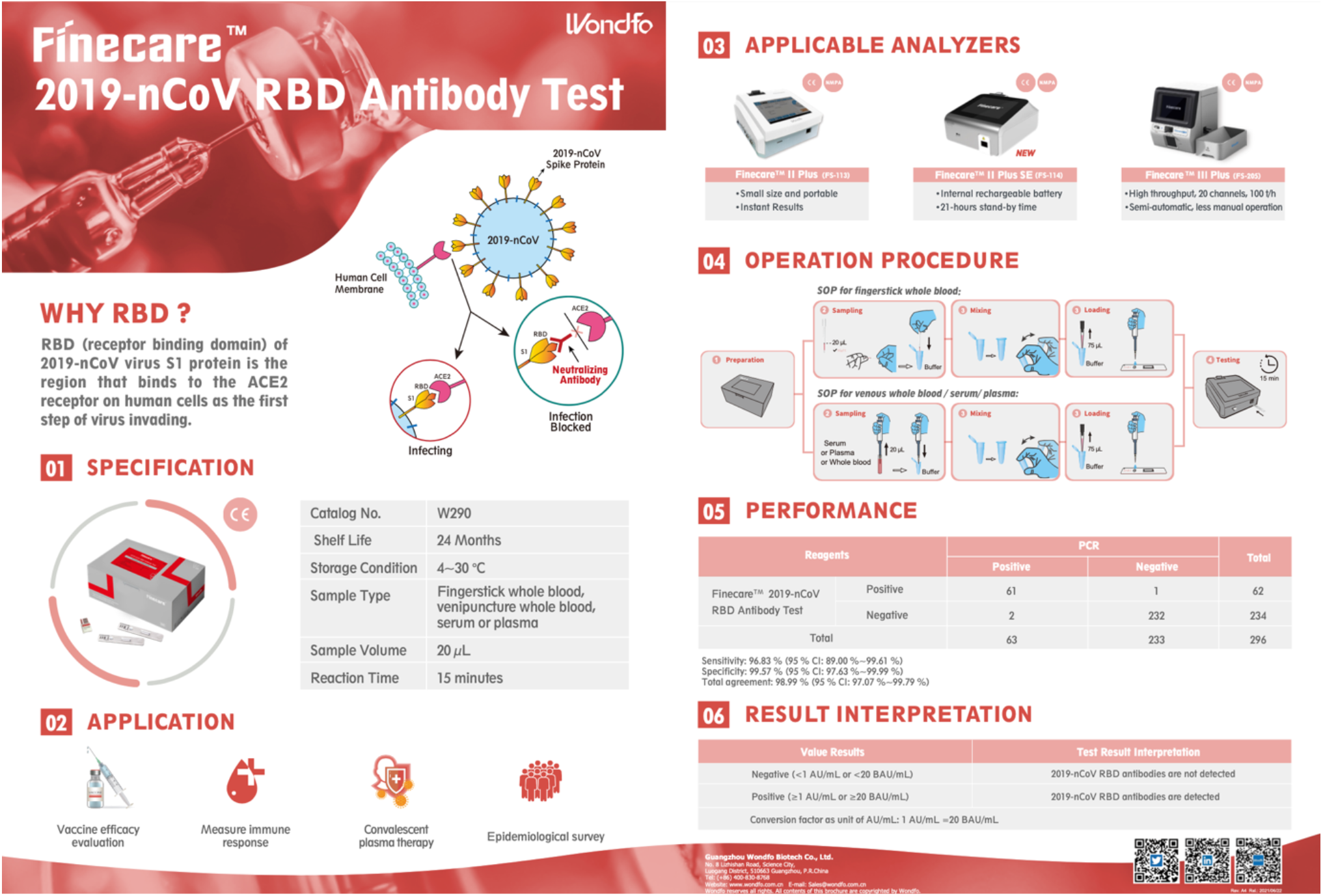
overview of FinecareTM2019-nCoV S-RBD lateral flow assay principle.

